# Filovirus Seroreactivity Beyond Virus-Specific Outbreak Regions in the Democratic Republic of the Congo, 2015-2023

**DOI:** 10.64898/2026.10.20.26366019

**Authors:** Sydney Merritt, Jean Paul Kompany, Patrick Mukadi, Megan Halbrook, Kamy Musene, Michael Beya, Handdy Kalengi, Merly Tambu, J. Daniel Kelly, Aquena H. Ball, Albert To, Daniel Mukadi-Bamuleka, Matthias Mossoko, Hannah L. Wallace, Teri Ann Wong, Reena H. Doshi, David McIlwain, Christopher W. Woods, Bradly P. Nicholson, Lisa E. Hensley, Jason Kindrachuk, Axel T. Lehrer, Placide Mbala-Kingebeni, Nicole A. Hoff, Anne W. Rimoin

## Abstract

The ongoing Bundibugyo virus disease (BVD) outbreak in the Democratic Republic of the Congo (DRC) is the second-largest Ebola disease outbreak in recorded history, and only the second recorded BVD outbreak in the country. While the DRC has recorded 18 filovirus disease outbreaks, the historical frequency and geographic distribution of human filovirus exposures remain poorly understood. Using a multiplexed multi-filovirus immunoassay, we assessed antibody reactivity against antigens of filoviruses linked to human disease, Bundibugyo virus (BDBV), Ebola virus (EBOV), Marburg virus (MARV), and Sudan virus (SUDV), among eleven cohorts from seven provinces between 2015–2023 (n=5,852). Participants with median fluorescence intensity values exceeding established antigen-specific cutoffs were classified as seroreactive. Seroreactivity against glycoprotein (GP) targets of all four viruses were detected in every cohort, demonstrating evidence of filovirus or an antigenically related exposure well beyond known outbreak regions. Seroreactivity estimates varied by cohort site, with 60.8% of participants (n=175/288) in Mongala Province in April 2016 considered seroreactive against BDBV GP. In this cohort, 4.2%, 29.9%, and 34.7% of participants were seroreactive against EBOV, MARV and SUDV GP, respectively yet only 1.7% were seroreactive against all four GP antigens. In adjusted analyses, site was the primary predictor of seroreactivity, with odds for BDBV seroreactivity 9.1-fold (95%CI 7.0-11.9) and 9.5-fold (95%CI 6.9-13.2) higher in Mongala and Tshuapa provinces than in Kwilu. Similarly, the adjusted odds of MARV and SUDV GP seroreactivity were 4.5-fold and 7.1-fold higher in Mongala, and 3.7-fold and 6.1-fold higher in Tshuapa, than Kwilu. Among 280 individuals sampled longitudinally, BDBV GP detected seroconversion was comparable by site, with 12.8% in Kwilu and 10.4% in Mongala having detected seroreactivity three and five years after initial contact, respectively. Overall, these results indicate widespread, geographically heterogeneous filovirus seroreactivity across sampled cohorts. These findings may reflect zoonotic spillover events, leading to undetected human-to-human filovirus circulation, including paucisymptomatic or asymptomatic infections, or exposure to as-yet unidentified antigenically cross-reactive viruses. These data highlight gaps in understanding the historical and geographic distribution of filovirus exposure in the DRC and support further prospective serological, epidemiological, and molecular investigations, especially given the current outbreak.

## INTRODUCTION

The Democratic Republic of the Congo (DRC) has documented 18 filovirus disease (FVD) outbreaks, more than any other country, since the first recognized emergence of *Orthoebolavirus zairense* (Ebola virus; EBOV) in 1976 near Yambuku, Mongala Province (1). Although most FVD outbreaks in DRC have been Ebola virus disease (EVD), both *Orthoebolavirus* and *Orthomarburgvirus* species have caused outbreaks in the DRC, reflecting the complicated and overlapping geographic distribution of filoviruses in the country, and Central Africa more broadly. This includes the re-emergence of *Orthoebolavirus bundibugyoense* (Bundibugyo virus; BDBV) identified on 15 May 2026 in Ituri Province, DRC, which at this time continues to be a public health emergency of international concern and the fastest growing outbreak recorded to date (2).

BDBV was first documented in DRC during a 2012 outbreak centered in Isiro, in what is now Haut-Uélé Province (3), five years after the virus was first identified during the 2007 outbreak in Bundibugyo District, Uganda, near the border with eastern DRC (4). In 1998, Marburg virus (MARV), was identified as the causative agent of a two-year Marburg virus disease (MVD) outbreak in the Watsa/Durba region also in the Haut Uele Province (5), primarily among gold miners. In contrast, no known Sudan virus disease (SVD) outbreaks have been documented in the DRC despite several recurrent SVD outbreaks in neighboring Uganda (1, 6).

Ebola and Marburg disease outbreaks typically follow a zoonotic spillover event from infected wildlife to humans (7, 8). Epidemiological investigations have shown that secondary human-to-human transmission can go undetected due to non-specific clinical presentations, suspect mild or asymptomatic infections, barriers to seeking healthcare, patterns of healthcare use, and limited surveillance, particularly in rural and remote settings, which have been historically under-resourced in Central Africa. Thus, the true geographic distribution and human exposure frequency for filoviruses within these regions remains uncertain.

Serological studies provide a means of reconstructing patterns of filovirus exposure over time that cannot be captured through acute illness disease surveillance alone (9). Antibody responses from FVD survivors can persist for decades following infection (10, 11), allowing specimens to provide insight into the historical and geographical distribution of exposure. However, interpretation of seroreactivity alone as an indicator of filovirus exposures in DRC can be further complicated given the potential for antigenic cross-reactivity with undocumented pathogens circulating in the region, as well as the possibility of waning humoral immunity which can vary with time since exposure and antibody response type. Thus, multiplexed immunoassays can help characterize seroreactivity across related and unrelated pathogens through the simultaneous detection of multiple antigens (12).

Understanding the historical distribution of filovirus exposure is increasingly important as outbreaks occur in new or unexpected locations. The unprecedented scale and geographic extent of the ongoing Bundibugyo virus disease (BVD) outbreak in the DRC where, as of October 18, 2026, 9,552 cases have been confirmed across 66 health zones of eight provinces (13, 14), further emphasize the critical need for surveillance. Of these cases, 4,602 deaths have been recorded (CFR = 48.2%). Moreover, the enormity of the current BVD outbreak in Northern DRC, as well as the lack of BVD-specific medical countermeasures, raises important questions regarding the true extent of filovirus circulation in the region, and potential incomplete detection of previous outbreaks. Here, we retrospectively assessed filovirus seroreactivity to BDBV, EBOV, MARV, and SUDV across twelve unique cohorts in seven provinces between 2015 and 2023 and investigated demographic, behavioral, occupational, and geographic factors associated with filovirus seroreactivity.

## METHODS

### Study Site Selection

Study sites were purposively selected in areas with previously documented FVD outbreaks, primarily focusing on past EVD outbreak locations **(Figure 1)**, which included Yambuku, Mongala Province (EBOV, 1976), Kikwit, Kwilu Province (EBOV, 1995), Boende, Tshuapa Province (EBOV, 2014), Mbandaka, Equateur Province (EBOV, 2018 and 2022), Beni, North Kivu Province (EBOV, 2018-2020, 2021, 2022) (1), and Watsa/Durba, Haut Uele Province (MARV, 1998-2000) (5). Kinshasa was also included as a recruitment site despite having not previously documented any FVD outbreaks, and was considered as a control population with likely limited exposure.

**Figure 1.**
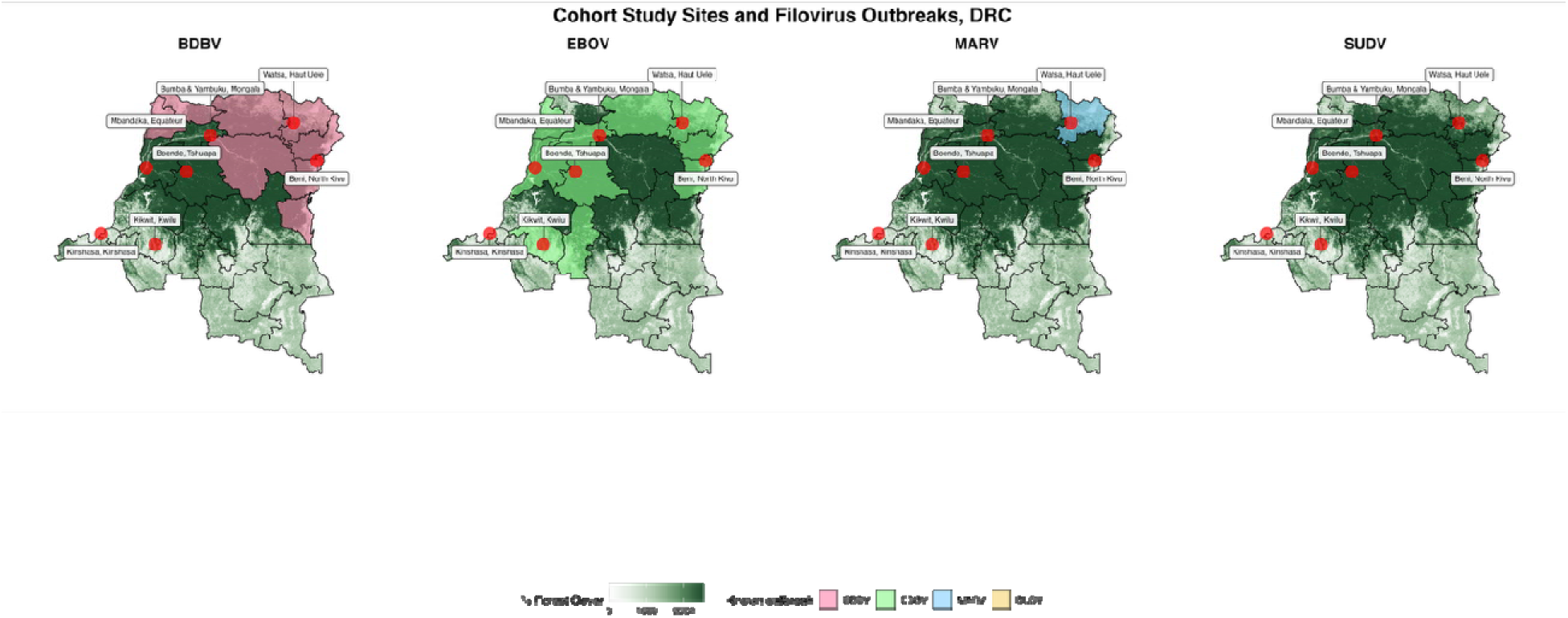
Cohort Recruitment Sites and Historic Filovirus Outbreaks in the Democratic Republic of the Congo. Provinces with confirmed outbreaks of BDBV, EBOV, and MARV are highlighted in pink, green, and blue; percent forest cover is indicated by green shading. No SUDV outbreaks have been reported in the DRC. Grey-shaded provinces indicate where cohorts included in this study were recruited. As of 18 October 2026, eight provinces have reported BDBV cases, with only Haut Uele reporting cases in both the 2012 and 2026 outbreaks.

Field collections were conducted between 2015 and 2023. Collection periods were not specifically scheduled to coincide with the rainy or dry season, yet the majority of cohort recruitment and sample collection was conducted during the dry season **(Supplementary Table 1).** While repeat cohort collections at the same site were not designed to be longitudinal collections of the same individuals, 280 individuals were sampled twice in Kikwit (n = 203, with ∼3 years in between collections) and Yambuku/Bumba (n = 77, ∼5 years). Only the most recent collection was included in presented tables and regression analyses, unless otherwise noted.

### Facility and Participant Eligibility and Selection

Within each study site, health facilities were selected by convenience from facilities located in health zones affected by previous FVD outbreaks. A list of all facilities (general hospitals, health centers and health posts) and an estimated number of employees at each site were provided by the health zone offices. However, due to logistical constraints such as distance and availability of health facility staff, not all facilities could be visited within every selected health zone area.

Eligible participants included known FVD survivors, close contacts and contacts of contacts of individuals with FVD, health-care workers, traditional healers, and other residents of the study area. When applicable, survivor populations were the primary target for enrollment, followed by healthcare workers and then other residents, contacts and family members of FVD survivors. There was no set recruitment target in each site, as populations in each site varied significantly. Attempts were made to contact all known FVD survivors in the area to inform them of the study and determine their interest in participating. Survivors were identified using Ministry of Health records and through local survivor organizations, including the Kikwit Ebola Survivors Association (15). Close contacts and contacts of contacts were identified through survivor referral, and specifically through the Ebola Family Association in Kikwit, Kwilu province. Individuals aged one year or older were eligible to participate if they belonged to one of the key populations recruited, and were at a selected site at the time of enrollment. Persons presenting with fever or considered unwell by a study physician at the time of enrollment were excluded.

All eligible HCWs with direct or indirect patient contact (16) were invited to participate if they worked at a selected health facility and were available at the time of sampling. Collection periods at each study site were typically one day, however, in the case of general hospitals and larger facilities, field collection teams (interviewers, phlebotomists, supervisors) may have enrolled participants for multiple days. Traditional healers were identified and recruited with the assistance of community health volunteers (“relais communautaires”) and were invited to visit selected health facilities closest to them during the study period. Other community residents who heard about the study either through community health volunteers, survivors, contacts, or HCWs were also eligible to participate if they were present at the selected health facility on a day a study team was enrolling participants. In some areas, participants from several nearby facilities were asked to congregate at a single selected facility for enrollment and specimen collection, rather than having study teams travel to multiple smaller sites.

Sampling in Tshuapa in September and November 2015 and in Mongala in January and April 2016 was unique in that these sites were the first field deployments of the larger study, when recruitment procedures and field logistics were being established. Initial visits at these two sites therefore focused primarily on community sensitization, establishing local study procedures, and assessing facility capacity for sample collection and storage, although some participant recruitment and sample collection also occurred. Subsequent visits focused on larger-scale recruitment and accounted for the majority of participants enrolled at these sites. Individuals enrolled in the initial scoping missions were not eligible to participate in the following larger recruitment visits that same year.

### Study procedures

All enrollment and study procedures were conducted at the selected health facilities. Parents or guardians provided written consent for minors who provided blood samples, and interview responses on behalf of the participating and assenting child. After consenting, participants were interviewed by trained local interviewers in French or their preferred local language (Lingala, Kikongo, or Swahili). Participants were provided with a transportation reimbursement based on local costs ranging between approximately ∼3 USD (rural locations) and ∼ 5 USD (Kinshasa) following study completion.

Interview questions included demographic characteristics, self-reported symptoms within the past six months, self-reported filovirus exposure, such as involvement in known outbreaks, vaccination, environmental, behavioural, and occupational exposure. Electronic survey records were hosted on secure servers–either ODKCollect (Get ODK Inc.) or SurveyCTO (Dobility Inc., Waltham MA).

Following the interview, anthropometric measures including height, weight, pulse and blood pressure were collected. All adult participants provided a ∼9 mL venous blood sample, collected by a trained phlebotomist on location. For minors who weighed less than 5 kilograms, no more than 1mL per kilogram was collected, however, there were no participants met this criterion for reduced blood draws. Blood samples were processed within 6 hours of collection at a dedicated field laboratory station, and serum was separated into 1 mL aliquots and stored in the field at -20°C. Samples were transported to the central laboratory in Kinshasa at -20°C and stored at -80°C until serological testing. Prior to testing, all biologic samples were heat-inactivated at 56°C for 30 minutes.

### Ethical Considerations

All study activities were approved by the Institutional Review Boards of the University of California, Los Angeles (IRB#15–0333, IRB#16–1346, IRB#20–1091) and the Kinshasa School of Public Health, University of Kinshasa (ESP/CE/038/2015, ESP/CE/022/2017, ESP/CE/142/2020). Participants provided written informed consent, or assent for minors, on a form which was signed or marked in agreement prior to commencement of enrollment activities.

### Serologic Evaluation

All serum samples were tested for antibody reactivity using a custom filovirus bead-based multiplexed immunological assay at the Institut National de Recherche Biomédicale (INRB) in Kinshasa. This multiplexed immunoassay had previously been assessed for result comparability between laboratory sites at the University of Hawaii and the INRB (12). In brief, magnetic microspheres (Luminex, Austin, TX) were covalently coupled with the following immunoaffinity purified antigens: BDBV glycoprotein (GP), EBOV GP, SUDV GP, MARV GP, EBOV nucleoprotein (NP), EBOV matrix protein VP40 and MARV VP40 (17). Bovine serum albumin (BSA)-coupled microspheres were included to control for non-specific binding. Samples were diluted to 1:100 and tested in duplicate, with mean MFI values presented for each target. All antigen-specific MFI output values were subsequently BSA-adjusted by subtracting detected anti-BSA MFI for each sample to eliminate possible unspecific protein-binding background by serum immunoglobulin G antibodies. Additional assay-specific details have been previously described (12). All testing was conducted between 2021 and 2023, with samples randomly pulled to minimize any batch effects in the presentation of serologic data.

Binary seroreactivity cut-offs were determined for each antigen by calculating the mean BSA-adjusted MFI value plus three standard deviations of a population of filovirus-naive individuals derived from the Kinshasa 2017 cohort (n = 379). This approach, and all antigen-specific cut offs, have been previously described in detail (18).

### Statistical Analyses

In total, 6,137 participants were enrolled and 5,852 (95.4%) were retained for final analysis. Only complete-case participants were retained for analysis, where questionnaire and serologic data were both available (n=285 excluded). Sensitivity analysis indicated that missingness of serologic and demographic data was completely at random across cohorts, bar serologic testing for reactivity against EBOV NP, due to antigen availability.

Descriptive statistics of categorical and binary variables are presented as counts and percentages, unless otherwise noted. We present seroreactivity to all four filovirus GP targets, as well as EBOV VP40, MARV VP40 and EBOV NP (where available), stratified by selected demographic characteristics, EBOV or MARV survivor status, and self-reported exposure in a past FVD outbreak. Given the non-random sampling approach with specific recruitment of known EBOV and MARV survivors, seroreactivity rates were subsequently calculated among all cohorts after excluding known survivors.

Multivariable logistic regression was employed to identify predictors of seroreactivity to all target antigens with adjustment for participant age, sex, and occupation. Variables of interest included: self-reported EBOV vaccine receipt, educational attainment as a proxy variable for socioeconomic status, province of recruitment (site), age, and sex. Time since last known outbreak was not included in the regression approach given the collinearity with sampling site, and missingness/no known outbreak in Kinshasa. Occupation was collapsed to fewer categories to account for small cell counts; “occupation - other” includes students, drivers, teachers, politicians, and non-healthcare staff, among others. Farmers, livestock farmers, and hunters were classified separately to assess potential workplace zoonotic exposures and associations with detected seroreactivity. Reported FVD exposure was also included as a key variable in all analyses, as participant recruitment was not a random community sample. Self-reported involvement in an FVD outbreak was collapsed into a categorical variable including reported close contact and/or employment during an outbreak. Kikwit, Kwilu province was specifically chosen as the reference group for the variable “site” to minimize potential over-inflation of reported odds ratios had Kinshasa been selected as the reference group—a subset of filovirus-naive individuals recruited in Kinshasa were the same individuals used to determine seroreactivity thresholds.

Among HCWs only, (n = 3,643) reported patient-contact and type (formal or informal) were classified as direct, indirect, unlikely or unknown, as has been previously reported (16). Regression analyses were conducted among HCW only to evaluate potential associations between HCW classification and seroreactivity.

To account for multiple hypothesis testing, all presented p-values are adjusted using the Benjamini-Hochberg procedure (minimizing false discovery rate). All analysis and figure generation was conducted in RStudio (R version 4.6.0, R Core Team, 2026).

### Ecological Classification

We used a suite of 10 geographic and environmental variables to characterize each sampling site and to compare the conditions under which seroreactive individuals were identified. Variables included three spatial variables (latitude, longitude, altitude), as well as seven remote-sensed environmental variables derived from spaceborne satellites. These variables captured the concentration of green leaf vegetation (MODIS derived NDVI-Normalized Difference Vegetation Index (19), the percentage of tree canopy cover (VCF, (20)), and surface moisture and leaf water content as monthly composites (mean) of global Quick Scatterometer (QSCAT, (21)). Together, we assessed the relative greenness, moisture, and tree cover of each site, and categorized these metrics into three major classes: Primary Forest, Secondary Forest, and Urban. We then assigned each site to one of these classifications and verified from expert opinions the validity of these categorizations.

## RESULTS

Across all eleven cohorts, 5,852 individuals were retained for this analysis and had complete data and results from serologic testing **(Table 1)**. More male participants recruited at all sites, except for Kinshasa and Kikwit. Moreover, HCWs comprised the majority of participants overall (62.3%). Among HCWs (n = 3,643), 94.5% were classified as formal HCW (physicians, nurses, working in a traditional health care setting) and 65.6% classified as likely having direct contact with patients **(Supplementary Table 2,** (16)). Across all cohorts, 20.6% of participants reported involvement in a past FVD outbreak, whether as a close contact, or having worked in an outbreak. Overall, 54 known survivors of filovirus infection (0.9%) were recruited, including 49 known EBOV survivors of the Yambuku, Kikwit, and Boende outbreaks, and 5 known MARV survivors of the Watsa/Durba outbreak. In Watsa/Durba, Equateur, and Mongala Provinces, 94 participants (1.6%) reported receiving an EBOV vaccine, rVSV-ZEBOV-GP, which was offered under a compassionate use/expanded access protocol during the 2018-2020 EBOV outbreak in Beni, North Kivu Province or the 2018 Mbandaka, Equateur (22)

**Table 1.**
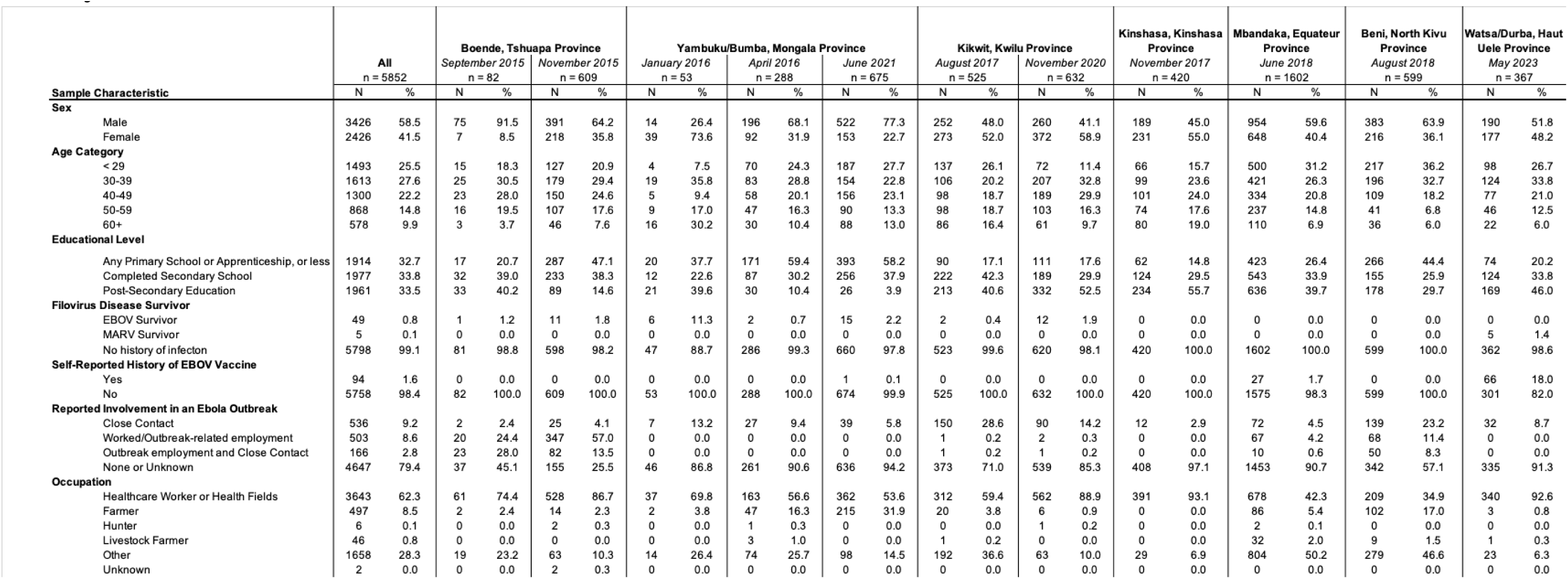
Cohort Demographic Characteristics (n = 5852). Sample characteristics are tabulated by collection site (province) and month, year of recruitment.

Overall, 1,947 participants (33.2%) displayed seroreactivity to any GP antigen, with BDBV GP having the highest overall seroreactivity (24.3%) across all cohorts. Anti-BDBV antibody seroreactivity was highest in Tshuapa, November 2015, and Mongala, April 2016 with 56.3% and 60.8% of individuals with detected seroreactivity, respectively **(Table 2**, **Figure 2)**. Conversely, across all cohorts, EBOV GP, VP40, and NP seroreactivity never exceeded 16.6%. For MARV and SUDV GP targets, the highest seroreactivity was also detected in Mongala, April 2016, with 29.9% and 34.7% of individuals classified as having seroreactivity, respectively. Only 41 individuals (0.7%) were classified as seroreactive to all four GP targets.

**Figure 2.**
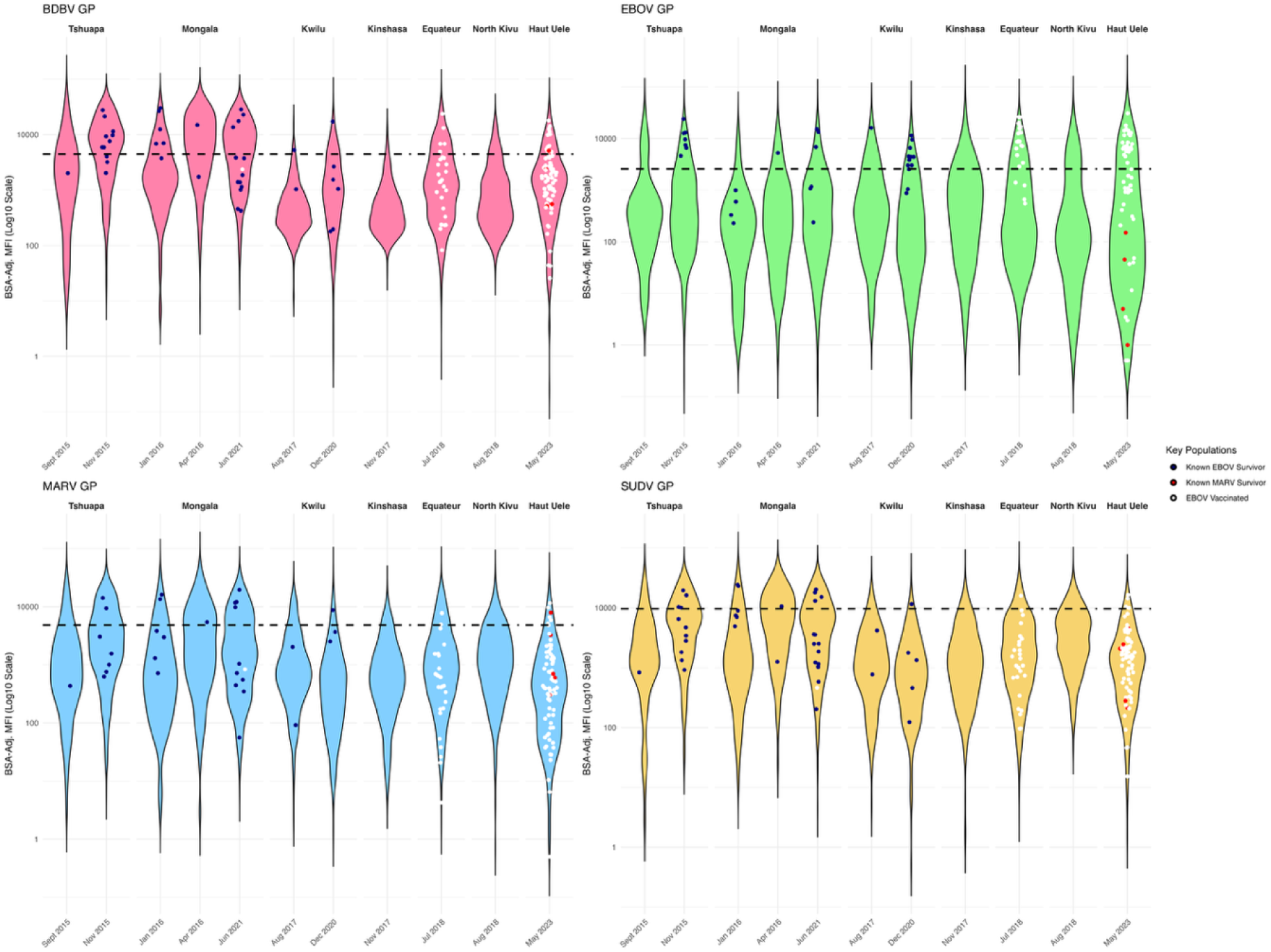
Violin Plots of Relative Median Fluorescence Intensity (MFI) Reactivity to Filovirus Glycoproteins by Province. Known survivors of EBOV or MARV as identified by Ministry of Health records are highlighted as blue and red points, respectively. Self-reported EBOV vaccine recipients are identified as white points. Antigen-specific binary seroreactivity cut offs are indicated by horizontal dashed lines

**Table 2.**
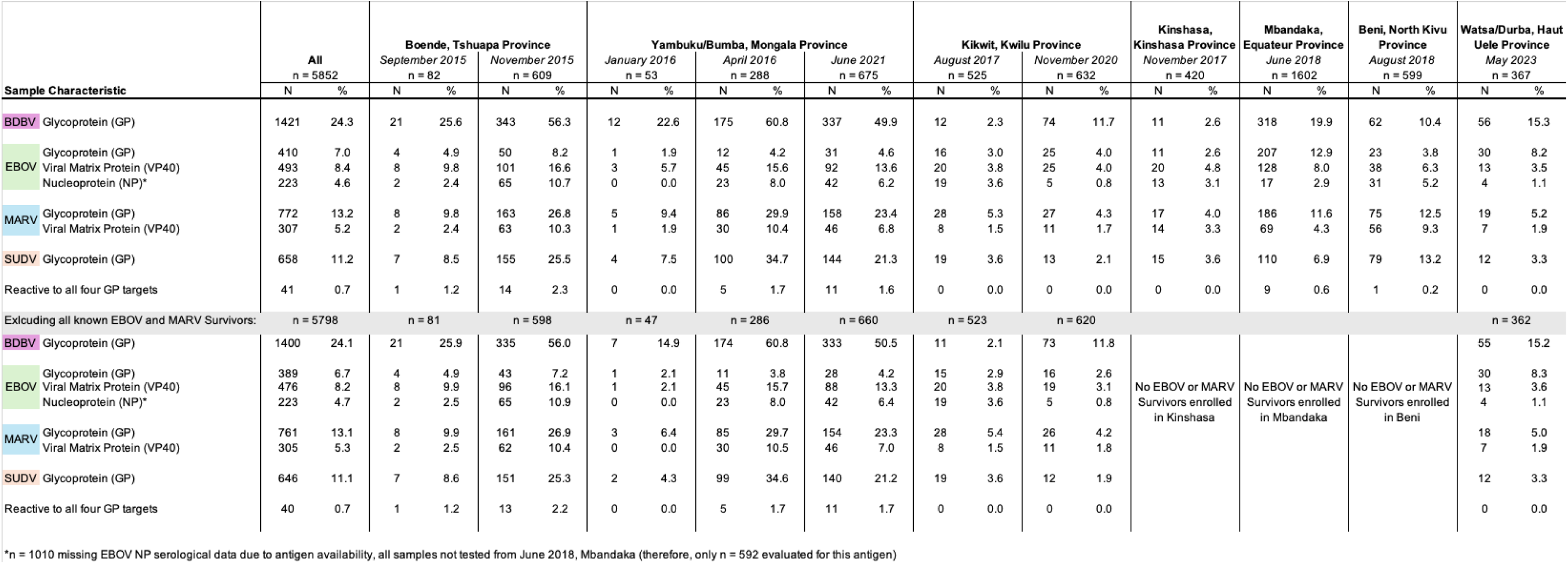
Filovirus Seroreactivity by Cohort. Percent seroreactivity is presented for all filovirus antigen targets included in the multiplexed assay by cohort site and collection visit. Binary seroreactivity is also presented following the exclusion of known MARV and EBOV survivors.

When excluding known FVD survivors, trends in seroreactivity against BDBV, MARV, and SUDV were largely unchanged with 24.2% of participants seroreactive against BDBV GP, 6.7% seroreactive against EBOV GP, 13.1% seroreactive against MARV GP, and 11.1% seroreactive against SUDV GP.

When assessing seroreactivity to individual and all possible combinations of GP targets **(Supplementary Figure 1)**, of the 1421 individuals seroreactive to BDBV GP, 711 (50.0%) were uniquely seroreactive to this target alone. An additional 27.9% (n=363/1421) of those with detected BDBV seroreactivity were also seroreactive to MARV and SUDV GP.

Adjusting for participant age, sex, reported FVD exposure, survivor status, education, occupation, and self-reported EBOV vaccination, the odds of BDBV GP seroreactivity in Tshuapa, Mongala, Equateur, and Haut-Uele provinces were 9.5-fold (95% CI: 6.9-13.2), 9.1-fold (95% CI: 7.0-11.9), 2.9-fold (95% CI: 2.2-3.8), and 1.9-fold (95% CI: 1.3-2.8) times the odds of BDBV GP seroreactivity in Kwilu, respectively **(Table 3**, **Figure 3)**. Notably, marked increases in the adjusted odds ratio (aOR) of seroreactivity against EBOV GP were only observed in Equateur (aOR = 3.9, 95%CI: 2.7-5.7). The adjusted odds of MARV GP and SUDV GP seroreactivity were significantly associated with site location in Equateur, Mongala, North Kivu and Tshuapa provinces.

**Table 3.**
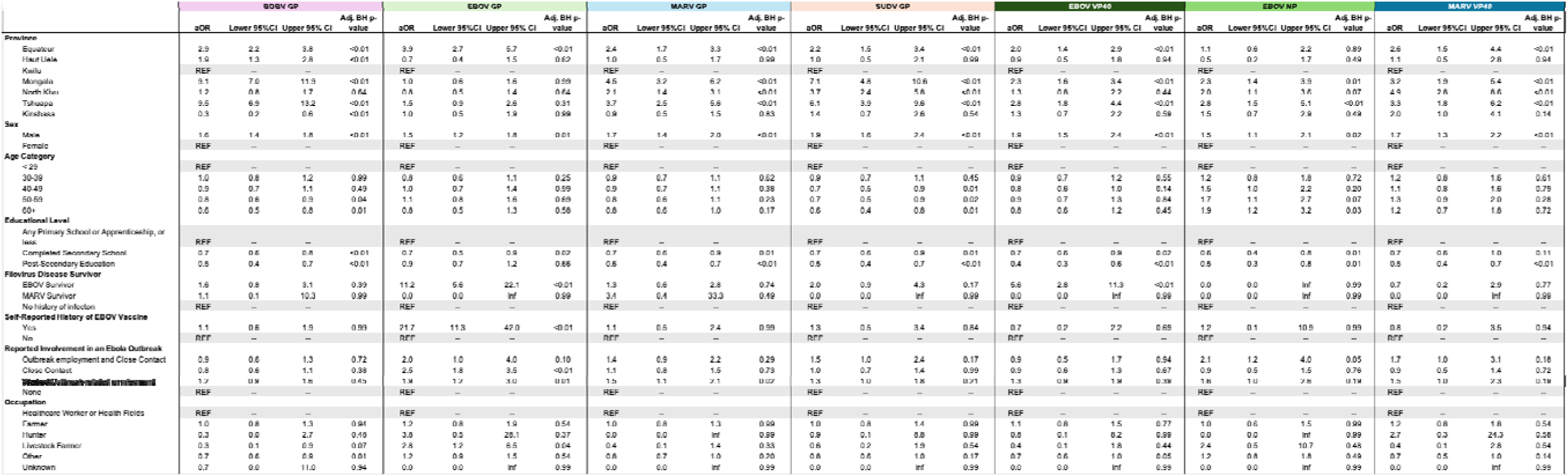
Adjusted Odds of Filovirus Seroreactivity. Adjusted odds of binary seroreactivity against BDBV, EBOV, MARV and SUDV GP, as well as EBOV VP40, EBOV NP and MARV VP40, are presented after adjusting for collection site (province), participant sex, age category, educational attainment, study enrollment criteria, occupation and self-reported EBOV vaccination status. Reference groups are indicated in grey. All presented p-values have been adjusted using the Benjamini-Hochberg (BH) to minimize false discovery rates.

**Figure 3.**
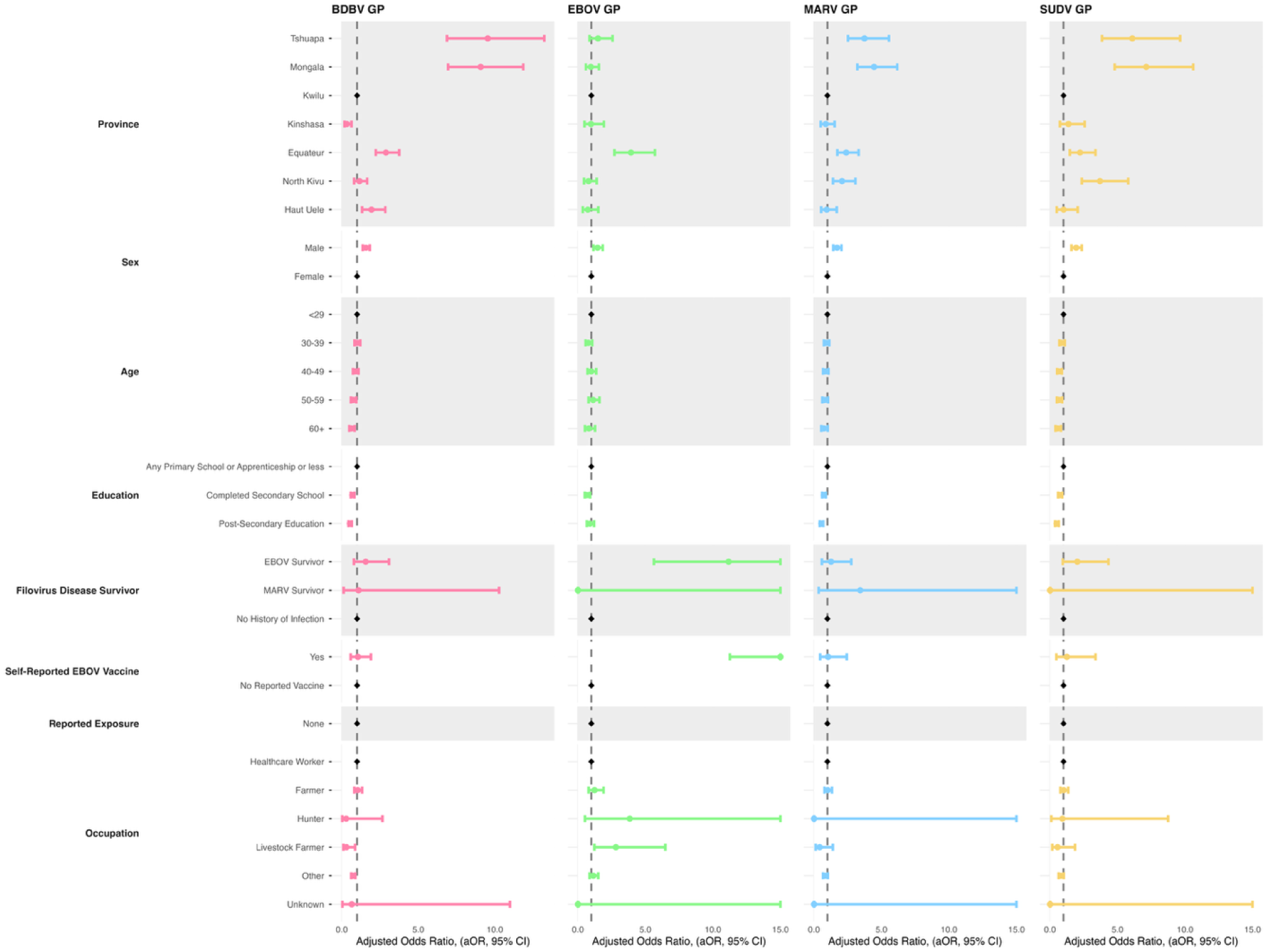
Forest Plots of Adjusted Odds Ratios of Seroreactivity to Filovirus Glycoproteins. Point estimates and 95% Confidence Intervals are denoted for each glycoprotein target by point and band width.

Multiple logistic regression analyses indicate that for all filovirus GP targets, the adjusted odds of seroreactivity are higher among male respondents, and those participants with a post-secondary education have reduced odds of seroreactivity when compared to those with primary schooling or no formal education. By age category, the odds of seroreactivity against BDBV GP were reduced in an almost stepwise manner with increasing age; when compared to those participants aged under 29 years old, older participants (age 60+) were less likely to be classified as seroreactive (aOR: 0.6, 95% CI: 0.5-0.8). Among known EVD survivors, the adjusted odds of seroreactivity against EBOV GP were 11.2-fold higher (95% CI: 5.6-22.1). Lastly, among those participants who reported receiving an EBOV vaccine (n = 94), the adjusted odds of EBOV seroreactivity are significantly increased (aOR: 21.7, 95% CI: 11.3-42.0), yet no parallel significant increases in the odds of seroreactivity to BDBV, MARV or SUDV GP were observed.

Moreover, the adjusted odds of seroreactivity against EBOV VP40 were not significantly increased (aOR = 0.7, 95%CI: 0.2-2.2) among those self-reporting an EBOV vaccine, yet were significantly higher among EBOV survivors (aOR = 5.6, 95%CI: 2.8-11.3). In Tshuapa, Equateur, and Mongala provinces, the adjusted odds of seroreactivity against EBOV VP40 were also elevated, but to a lesser extent than any GP target (Tshuapa aOR = 2.8, 95%CI: 1.8-4.4, Equateur aOR = 2.0, 95%CI: 1.4-2.9, Mongala aOR = 2.3, 95%CI: 1.6-3.4). When repeating analysis after excluding all known FVD survivors (n = 5,798), trends in the adjusted odds of seroreactivity by site, age and occupation remain unchanged **(Supplementary Table 3)**. Similarly, when additionally excluding self-reported EBOV vaccine recipients, adjusted odds ratios remain stable (n = 5,704).

Among HCW only, the adjusted odds of BDBV GP seroreactivity were significantly increased among “informal” HCW, in comparison to formal HCW (aOR = 1.9, 95%CI: 1.1-3.3). This increased odds of seroreactivity was not observed for EBOV, MARV or SUDV GP **(Supplementary Table 4).**

While not a primary study objective, nested longitudinal collection of 280 individuals allowed for the detection of seroconversion events **(Table 4)**. Overall seroreactivity rates against all four GP targets were lower in Kwilu than in Mongala, yet detected seroconversion over time to BDBV GP was comparable, with 12.8% of individuals in Kikwit and 10.4% of individuals in Yambuku with detected seroreactivity three and five years later, respectively. However, this longitudinal capture also indicated seroreversion—in Yambuku, 22.1%, 24.7% and 22.1% of participants reactive at first visit were no longer reactive five years later against BDBV, MARV and SUDV GP, respectively.

**Table 4.** Detected Changes in Seroreactivity Among Longitudinal Sub-cohorts (n = 280).

| Population | Time between visits | Longitudinal Serostatus | BDBV GP |  | EBOV GP |  | MARV GP |  | SUDV GP |  |
| --- | --- | --- | --- | --- | --- | --- | --- | --- | --- | --- |
|  |  |  | N | % | N | % | N | % | N | % |
| Kikwit, Kwilu (n = 203) | 3 Years | Seroconversion | 26 | 12.8 | 4 | 2.0 | 10 | 4.9 | 3 | 1.5 |
|  |  | Seroreversion | 9 | 4.4 | 7 | 3.4 | 7 | 3.4 | 1 | 0.5 |
|  |  | Stable Seronegative | 167 | 82.3 | 182 | 89.7 | 184 | 90.6 | 197 | 97.0 |
|  |  | Stable Seroreactive | 1 | 0.5 | 10 | 4.9 | 2 | 1.0 | 2 | 1.0 |
| Yambuku/Bumba, Mongala (n = 77) | 5 years | Seroconversion | 8 | 10.4 | 2 | 2.6 | 4 | 5.2 | 8 | 10.4 |
|  |  | Seroreversion | 17 | 22.1 | 5 | 6.5 | 19 | 24.7 | 17 | 22.1 |
|  |  | Stable Seronegative | 24 | 31.2 | 66 | 85.7 | 41 | 53.2 | 43 | 55.8 |
|  |  | Stable Seroreactive | 28 | 36.4 | 4 | 5.2 | 13 | 16.9 | 9 | 11.7 |

Unique sites, as identified by geographic coordinates collected at the time of sample collection, were classified by described ecological parameters as primary forest, secondary forest or urban settlements. Mongala, Tshuapa, and Equateur were classified as primary forest, Kwilu, Haut Uele, and North Kivu were classified as secondary forest, and Kinshasa was classified as urban **(Table 5).**

**Table 5.**
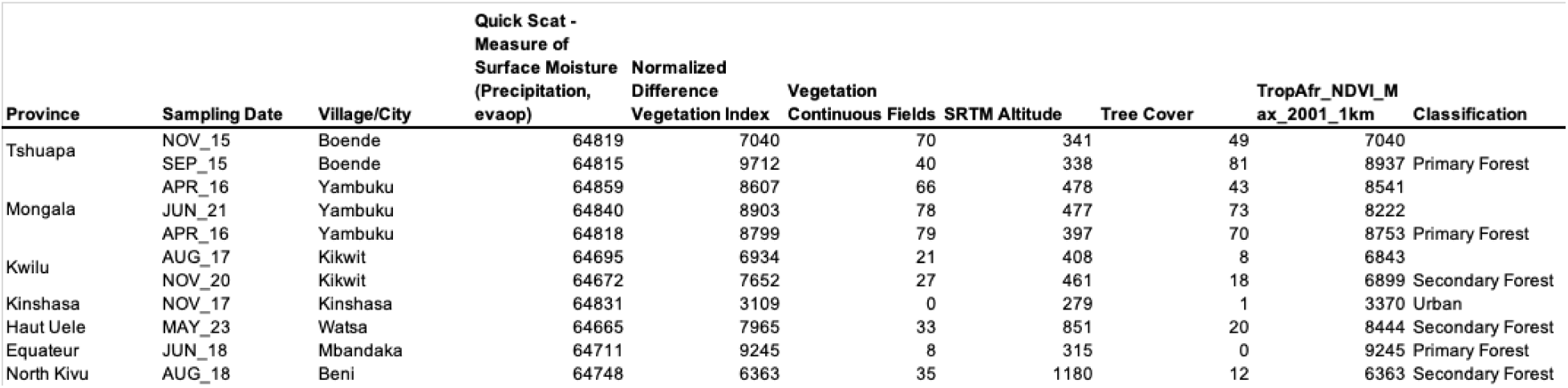
Ecological Classifications of Cohort Sites.

## DISCUSSION

This study provides a broad assessment of filovirus seroreactivity across geographically and epidemiologically diverse populations in DRC, including areas with and without previously documented filovirus outbreaks. In this study, we identified elevated seroreactivity against BDBV GP across diverse cohorts in seven provinces in DRC. Although cohorts are not population-representative, these data provide important insights into circulating filoviruses where, in provinces such as Mongala and Tshuapa, over half of the sampled population had a detectable antibody response against BDBV GP. Beyond BDBV, we also detected seroreactivity against MARV and SUDV, at sites with no history of reported MVD or SVD outbreaks.

Serologic evidence of filovirus exposure in populations with no history of infection has been documented previously. For example, in a study from Kasai Oriental province, 11% of 3415 individuals had detectable anti-EBOV antibodies (23). In the nearby Central African Republic (CAR), among a population of farmers and hunter-gatherers collected from 1992-1994, 5.3% and 2.4% were seroreactive against EBOV and MARV, respectively (24). More recently, evidence from Guinea has indicated seroreactivity rates of 4.9% and 4.2% against EBOV and BDBV among a population of wildlife-hunter communities sampled in 2017 (25). Similarly, a more regional retrospective study of bio banked serum samples collected from 1997-2012 documented reactivity against EBOV GP among individuals in the Republic of the Congo and the DRC (26), further emphasizing the likely underestimation of FVD circulation regionally. BDBV seroreactivity has also been reported, although rarely, in cohorts from Uganda outside recognized BVD outbreak areas (27). These observations support the possibility that BDBV or antigenically related viruses have circulated beyond locations where FVD outbreaks have been documented.

Taken together, these results may indicate there is ongoing circulation of filoviruses, or more broadly, viruses that share strong sequence similarities and common epitopes with BDBV, MARV and SUDV. Such circulation could remain undetected, particularly if infections are asymptomatic or paucisymptomatic, health-care seeking is limited or delayed, or transmission chains are small and end before an outbreak is recognized (28).

The elevated rates of seroreactivity against BDBV, EBOV and SUDV GPs among participants without known FVD diagnoses may be attributed to another antigenically similar, yet unidentified, virus. Although filoviruses are often associated with substantial morbidity and mortality, both Reston virus (RESTV) and Bombali virus (BOMV) are known *Orthoebolaviruses* that have not been associated with pathogenic disease in humans; however, neither virus has been documented in DRC. Given this, as well as the conserved structural features of the *Orthoebolavirus* GPs (29, 30), it is possible that infection with an unknown filovirus with mild or entirely asymptomatic disease could induce antibodies that cross-react with BDBV, EBOV or SUDV GP. Substantial serologic cross-reactivity has also been demonstrated among *Orthoebolaviruses*; cross-reactive IgG responses to heterologous ebolavirus antigens have been detected in convalescent sera from EBOV survivors (31), and isolated monoclonal antibodies from survivors of the 2007 BVD outbreak demonstrated potent neutralizing activity against multiple filovirus species (32). In sum, these findings support the possibility of exposure to an antigenically related filovirus which may play a role in the detected seroreactivity.

Moreover, the geographic range of filoviruses has been shown to be far more extensive than previously recognized with the identification of non-*orthoebolaviruses* or *orthomarburgviruses* in Europe and Asia (33, 34). While outside of the scope of the current study and available methodology, future research should evaluate potential underlying seroreactivity to all known filoviruses, including those that have not been known to cause human disease, as well as supporting additional surveillance efforts to identify other novel viruses. These findings can then guide more targeted epidemiologic, ecological, and virologic investigations, including surveillance and virus discovery efforts in areas where unrecognized filovirus circulation may be occurring.

In multivariable analyses, geographic location was strongly associated with the less common filoviruses, BDBV, MARV and SUDV GP seroreactivity, with higher seroreactivity among participants from Yambuku and Bumba in Mongala Province and Boende in Tshuapa Province. These sites were also classified as “primary forest” based on ecological characteristics, including normalized difference vegetation index, vegetation continuous fields, and tree cover. Conversely, sites such as Kikwit and Watsa, which are sites of past EBOV or MARV outbreaks, are located in notably less dense, or “secondary forest” ecological zones. Of note, no samples were collected from Isiro, Haut Uele province (2012 BDBV outbreak), yet this site is <300 kilometers from the Watsa/Durba region and has similar ecological characteristics. However, no increase in adjusted odds of anti-BDBV seroreactivity was observed, relative to other provinces such as Tshuapa and Mongala. Although ecological characteristics were not included in the regression models, this geographic pattern observed here supports the notion that forest environment could influence opportunities for zoonotic exposure. Additionally, all geographic classifications were determined with data from the date of sample collection, yet this may not reflect true forest cover at the time of the known outbreaks, which, in some cases, were up to forty years prior to sample collection. Further studies incorporating individual-level exposure data and environmental characteristics are needed to assess whether forest density or related ecological factors are directly associated with filovirus spillover and repeated human exposure.

Among those participants who self-reported receipt of an EBOV vaccine, no significant increase in the odds of BDBV, MARV or SUDV GP, seroreactivity was observed. Additionally, no observed increase in seroreactivity against EBOV VP40 and EBOV NP among vaccinated populations exemplifies the specificity of the assay for detecting vaccine-induced anti-EBOV GP antibodies. However, the absence of data on the date of vaccination, or baseline seroreactivity prior to vaccination limits interpretation of this finding and precludes conclusions regarding the ability of the rVSV-EBOV-GP vaccine to induce cross-reactive responses against BDBV GP. This vaccine was deployed under a compassionate use/expanded access protocol in the DRC first in 2018 in both Mbandaka and Beni, suggesting that all samples from participants in cohorts from 2021 and 2023 who reported vaccination were collected likely between two- and five-years post-vaccination, but this cannot be confirmed. Thus, the absence of detectable BDBV GP seroreactivity by the assay could reflect waning cross-reactive antibody responses over time rather than an absence of vaccine-induced cross-reactivity among these cohorts. Alternatively, of the 27 vaccinated individuals in Mbandaka, sample collection was likely within weeks of vaccine reception, further complicating interpretation of serologic results. This distinction is particularly relevant considering the growing interest in the potential use of the existing EVD vaccine as countermeasures in the current BVD outbreak (17, 35–38).

This study has several limitations. First, these data were collected from convenience samples at purposively selected sites with previous FVD outbreaks, with enrollment targeting known survivors, HCWs, and other individuals with potential previous FVD exposure. The majority of the samples were from HCW, given that previous research identified elevated levels of anti-EBOV antibodies among HCW cohorts in the DRC (16), the frequency of seroreactivity observed in this study should not be interpreted as population-level seroprevalence and may not be generalizable to the broader populations of the sampled provinces or the DRC as a whole.

Second, utilizing a multiplexed immunoassay approach provides multi-target reactivity estimates, yet serologic assessments are not without their limitations. Primarily, detected seroreactivity is not definitive evidence of antibody functionality or neutralization potential, as not all antibodies detected will mediate protection (39). Additionally, observed seroreactivity may instead reflect nonspecific antibody binding be due to an recent or concurrent infections, such as malaria, which is endemic in the DRC, and may induce subsequent systemic immune activation and inflammation (40). For BDBV GP seroreactivity, our findings in the Mbandaka 2018 cohort (19.9%) were elevated in comparison with 2.4% previously detected using a single-plex assay (41). While no data on current or past malaria infection was collected as a part of this study, evidence of false-positive serologic tests during acute malaria infection (40) should not be discounted as yet another element in interpretation of seroreactivity.

Furthermore, these cross-sectional data cannot be used to draw causal conclusions, given the single time-point collection. While BDBV seroconversion was detected among convenience longitudinal samples in Kikwit and Yambuku (n = 280), additional clinical data, medical records, and molecular testing would be needed to definitively suggest filovirus infection or exposure.

Finally, associations identified in the multivariable analyses should be interpreted with caution due to the potential for substantial residual and unmeasured confounding. Available covariates differed across cohorts, and potentially important factors could not be consistently accounted for, including zoonotic exposures and contact with wildlife, contact with patients or individuals with a similarly presenting illness, previous health-care seeking or medical care, occupational exposures, and other behavioural and environmental factors. The targeted recruitment strategy could also introduce selection bias and associations between measured characteristics that differ from those in the underlying population. Consequently, observed associations, particularly those with geographic location and occupation, should not be interpreted as causal or as evidence of specific routes of filovirus exposure.

Future work should investigate the role of population mobility in shaping patterns of seroreactivity, and potential undetected continued spread of filoviruses in the DRC (42). Prospective studies incorporating more detailed exposure histories are also needed to assess behavioural and occupational factors associated with seroreactivity, including PPE use among HCW, contact with individuals with filovirus-compatible illness, burial practices, healthcare seeking, and the frequency and nature of contact with wildlife. These should be conducted both in historical outbreak regions, as well as those with no documented FVD, as evidenced by detected seroreactivity.

To our knowledge, this study represents the largest retrospective assessment of filovirus seroreactivity in the DRC, including 5,852 individuals across seven provinces. The detection of BDBV, EBOV, MARV, and SUDV GP seroreactivity beyond locations with recognized outbreaks suggests that the documented geographic distribution of FVD outbreaks may not fully reflect patterns of filovirus exposure in the DRC. These findings support broader investigations of filovirus seroreactivity, including in areas without previously documented FVD outbreaks, and highlight the value of longitudinal serological studies for understanding the historical and geographic distribution of filoviruses. Such evidence could help refine understanding of populations and areas at risk, provide an estimate baseline reactivity for future vaccine studies, and inform preparedness for future filovirus emergence.

## Data Availability

Data underlying the findings of this study can be made available upon reasonable request to the corresponding author.

## ACKNOWLEGEMENTS

The authors would first like to thank all study participants for their time and contribution to this work, as well as the participating communities, health facilities, provincial health authorities, and the Ministry of Health of the Democratic Republic of the Congo for their support in conducting the study. We are especially grateful to the local interviewers, phlebotomists, community health volunteers, and other field staff who supported participant recruitment, data collection, and specimen collection across the study sites.

The authors would like to thank Dr. Ryan J. Harrigan for his thoughtful review and contributions to the ecological data. The authors would also like to thank Rachel Mutombe, Nathalie Kavira, and Carine Mpeti for their contributions to this study, and Cyrus Sinai for his review of this manuscript.

## SUPPLEMENTARY MATERIAL

**Supplementary Table 1.** Cohort recruitment and sampling.

| Cohort Site | Collection Dates |  |
| --- | --- | --- |
|  | Start Date | End Date |
| **Boende, Tshuapa Province** |  |  |
| Visit 1 | September 14, 2015 | September 17, 2015 |
| Visit 2 | November 16, 2015 | November 26, 2015 |
| **Yambuku/Bumba, Mongala Province** |  |  |
| Visit 1 | January 27, 2016 | January 28, 2016 |
| Visit 2 | April 23, 2016 | April 30, 2016 |
| Visit 3 | June 27, 2021 | July 16, 2021 |
| **Kikwit, Kwilu Province** |  |  |
| Visit 1 | August 18, 2017 | September 8, 2017 |
| Visit 2 | November 21, 2020 | November 28, 2020 |
| **Kinshasa, Kinshasa Province** |  |  |
| Visit 1 | November 23, 2017 | December 8, 2017 |
| **Mbandaka, Equateur Province** |  |  |
| Visit 1 | June 7, 2018 | July 24, 2018 |
| **Beni, North Kivu Province** |  |  |
| Visit 1 | August 15, 2018 | August 29, 2018 |
| **Watsa/Durba, Haut Uele Province** |  |  |
| Visit 1 | May 16, 2023 | May 31, 2023 |

**Supplementary Figure 1.**
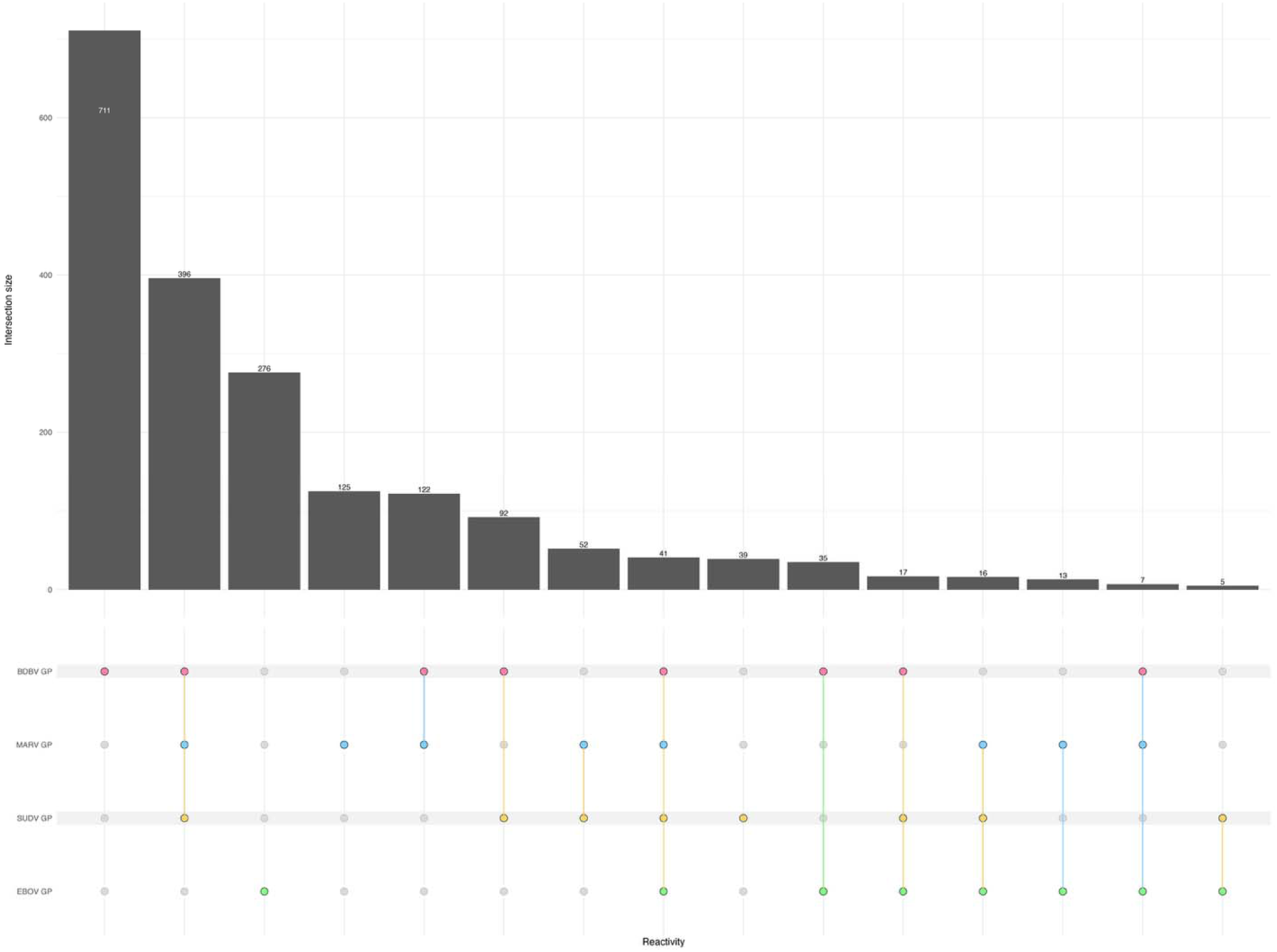
Upset Plot of Detected Multiple Filovirus GP Seroreactivity. While seroreactivity is binarized based on established cut offs, this plot demonstrates those individuals with detected seroreactivity to multiple GP targets at the time of sample collection.

**Supplementary Table 2.** Demographic Characteristics among Health Care Workers (HCW, n = 3,643).

| Sample Characteristic | All<br>n = 3643 |  | Boende, Tshuapa Province<br>September 2015<br>n = 61 |  | November 2015<br>n = 528 |  | Yambuku/Bumba, Mongala Province<br>January 2016<br>n = 37 |  | April 2016<br>n = 163 |  | June 2021<br>n = 362 |  | Kikwit, Kwilu Province<br>August 2017<br>n = 312 |  | November 2020<br>n = 562 |  | Kinshasa, Kinshasa Province<br>November 2017<br>n = 391 |  | Mbandaka, Equateur Province<br>June 2018<br>n = 678 |  | Beni, North Kivu Province<br>August 2018<br>n = 209 |  | Watsa/Durba, Haut Uele Province<br>May 2023<br>n = 340 |  |
| --- | --- | --- | --- | --- | --- | --- | --- | --- | --- | --- | --- | --- | --- | --- | --- | --- | --- | --- | --- | --- | --- | --- | --- | --- |
|  | N | % | N | % | N | % | N | % | N | % | N | % | N | % | N | % | N | % | N | % | N | % | N | % |
| Complete case healthcare workers only |  |  |  |  |  |  |  |  |  |  |  |  |  |  |  |  |  |  |  |  |  |  |  |  |
| **Healthcare Worker Type** |  |  |  |  |  |  |  |  |  |  |  |  |  |  |  |  |  |  |  |  |  |  |  |  |
| Formal | 3444 | 94.5 | 61 | 100.0 | 499 | 94.5 | 37 | 100.0 | 144 | 88.3 | 339 | 93.6 | 308 | 98.7 | 559 | 99.5 | 391 | 100.0 | 589 | 86.9 | 179 | 85.6 | 338 | 99.4 |
| Informal | 71 | 1.9 | 0 | 0.0 | 29 | 5.5 | 0 | 0.0 | 19 | 11.7 | 21 | 5.8 | 0 | 0.0 | 0 | 0.0 | 0 | 0.0 | 2 | 0.3 | 0 | 0.0 | 0 | 0.0 |
| Unknown | 128 | 3.5 | 0 | 0.0 | 0 | 0.0 | 0 | 0.0 | 0 | 0.0 | 2 | 0.6 | 4 | 1.3 | 3 | 0.5 | 0 | 0.0 | 87 | 12.8 | 30 | 14.4 | 2 | 0.6 |
| **Healthcare Worker Contact Type** |  |  |  |  |  |  |  |  |  |  |  |  |  |  |  |  |  |  |  |  |  |  |  |  |
| Direct | 2388 | 65.6 | 37 | 60.7 | 263 | 49.8 | 30 | 81.1 | 117 | 71.8 | 259 | 71.5 | 236 | 75.6 | 439 | 78.1 | 240 | 61.4 | 415 | 61.2 | 141 | 67.5 | 211 | 62.1 |
| Indirect | 774 | 21.2 | 22 | 36.1 | 197 | 37.3 | 6 | 16.2 | 35 | 21.5 | 66 | 18.2 | 50 | 16.0 | 78 | 13.9 | 86 | 22.0 | 124 | 18.3 | 35 | 16.7 | 75 | 22.1 |
| Unlikely | 353 | 9.7 | 2 | 3.3 | 68 | 12.9 | 1 | 2.7 | 11 | 6.7 | 35 | 9.7 | 22 | 7.1 | 42 | 7.5 | 65 | 16.6 | 52 | 7.7 | 3 | 1.4 | 52 | 15.3 |
| Unknown | 128 | 3.5 | 0 | 0.0 | 0 | 0.0 | 0 | 0.0 | 0 | 0.0 | 2 | 0.6 | 4 | 1.3 | 3 | 0.5 | 0 | 0.0 | 87 | 12.8 | 30 | 14.4 | 2 | 0.6 |

**Supplementary Table 3.**
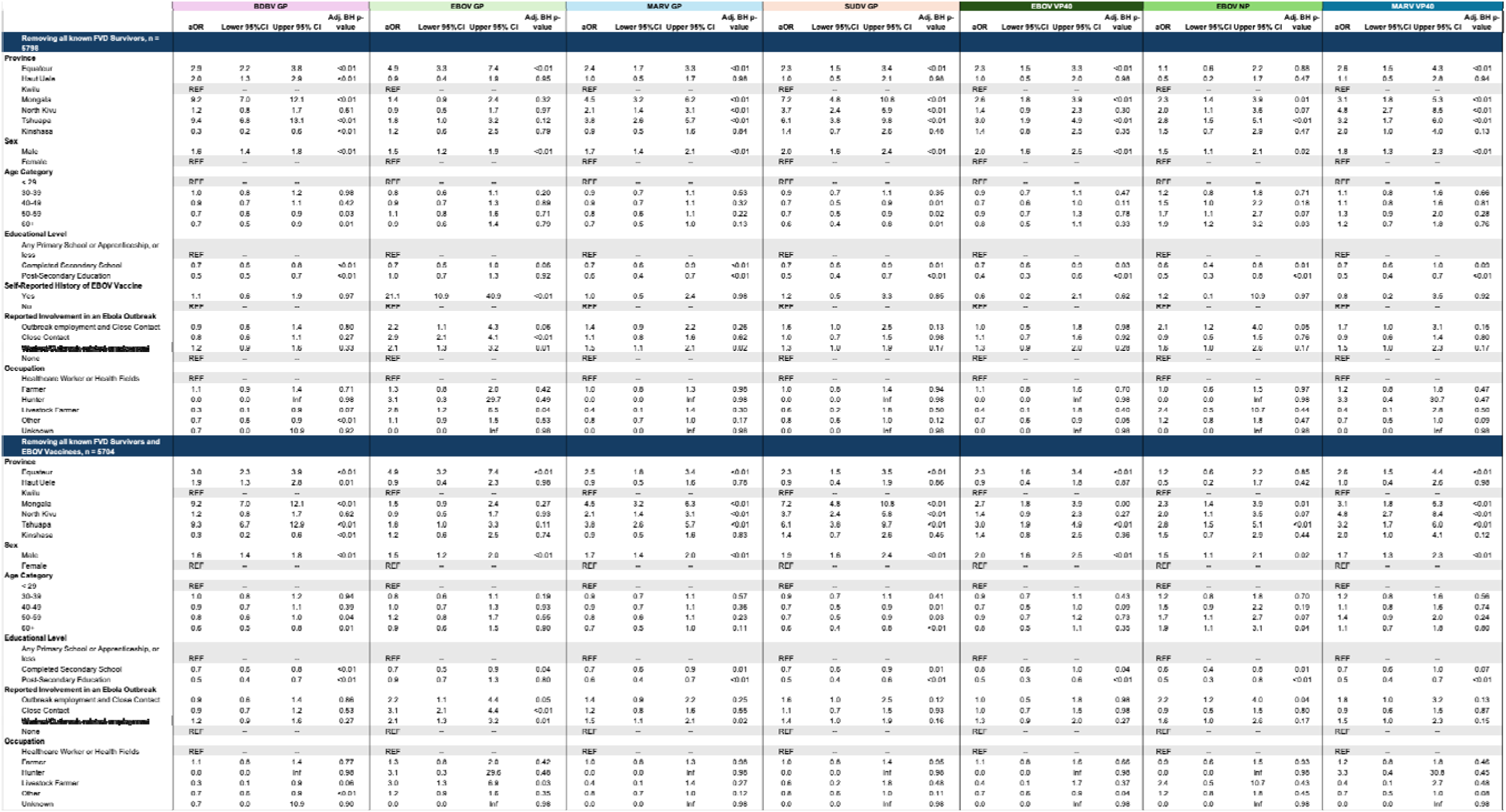
Adjusted Odds of Filovirus Seroreactivity Removing Known Survivors and Self-Reported EBOV Vaccine Recipients. Adjusted odds of binary seroreactivity against BDBV, EBOV, MARV and SUDV GP, as well as EBOV VP40, EBOV NP and MARV VP40, are presented after adjusting for collection site (province), participant sex, age category, educational attainment, study enrollment criteria, occupation and self-reported EBOV vaccination status. Reference groups are indicated in grey. All presented p-values have been adjusted using the Benjamini-Hochberg (BH) to minimize false discovery rates.

**Supplementary Table 4.**
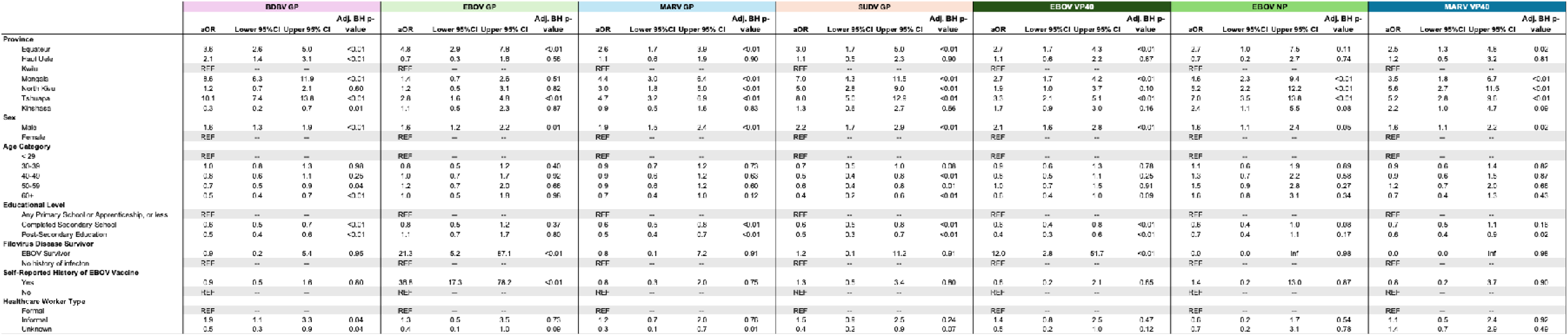
Adjusted Odds of Filovirus Seroreactivity among HCW Only.

## REFERENCES

1. Sullivan NJ. Bundibugyo virus disease in 2026—clinical and public health responses. New England Journal of Medicine. 2026;395(3):278–89.

2. Organization WH. Epidemic of Ebola Disease caused by Bundibugyo virus in the Democratic Republic of the Congo and Uganda determined a public health emergency of international concern 2026 [Available from: https://www.who.int/news/item/17-05-2026-epidemic-of-ebola-disease-in-the-democratic-republic-of-the-congo-and-uganda-determined-a-public-health-emergency-of-international-concern.

3. Kratz T, Roddy P, Tshomba Oloma A, Jeffs B, Pou Ciruelo D, de la Rosa O, et al. Ebola Virus Disease Outbreak in Isiro, Democratic Republic of the Congo, 2012: Signs and Symptoms, Management and Outcomes. PLOS ONE. 2015;10(6):e0129333.

4. Towner JS, Sealy TK, Khristova ML, Albariño CG, Conlan S, Reeder SA, et al. Newly discovered ebola virus associated with hemorrhagic fever outbreak in Uganda. PLoS pathogens. 2008;4(11):e1000212.

5. Bausch DG, Nichol ST, Muyembe-Tamfum JJ, Borchert M, Rollin PE, Sleurs H, et al. Marburg hemorrhagic fever associated with multiple genetic lineages of virus. New England Journal of Medicine. 2006;355(9):909–19.

6. Whitworth HS, Quintard G, Oyeneye V, Postler TS, Marini A, Zaric M, et al. Ebola virus and Sudan virus infection in humans: a comparison to inform vaccine research and development. Emerg Microbes Infect. 2026;15(1):2686465.

7. Rouquet P, Froment JM, Bermejo M, Kilbourn A, Karesh W, Reed P, et al. Wild animal mortality monitoring and human Ebola outbreaks, Gabon and Republic of Congo, 2001-2003. Emerg Infect Dis. 2005;11(2):283–90.

8. Organization WH. Ebola disease 2025 [updated 24 April 2025. Available from: https://www.who.int/news-room/fact-sheets/detail/ebola-disease.

9. Haselbeck AH, Im J, Prifti K, Marks F, Holm M, Zellweger RM. Serology as a Tool to Assess Infectious Disease Landscapes and Guide Public Health Policy. Pathogens. 2022;11(7).

10. Natesan M, Jensen SM, Keasey SL, Kamata T, Kuehne AI, Stonier SW, et al. Human Survivors of Disease Outbreaks Caused by Ebola or Marburg Virus Exhibit Cross-Reactive and Long-Lived Antibody Responses. Clin Vaccine Immunol. 2016;23(8):717–24.

11. Rimoin AW, Lu K, Bramble MS, Steffen I, Doshi RH, Hoff NA, et al. Ebola Virus Neutralizing Antibodies Detectable in Survivors of theYambuku, Zaire Outbreak 40 Years after Infection. J Infect Dis. 2018;217(2):223–31.

12. Smith OA, Merritt S, Kompany JP, Hoff NA, Wong TAS, Kamara V, et al. Multiplex pan-filovirus assay performance and reproducibility across varied geographical and resource settings. PLOS ONE. 2026;21(8):e0356257.

13. Organization WH. Ebola disease caused by Bundibugyo virus - Democratic Republic of the Congo Geneva2026 [Available from: https://www.who.int/emergencies/disease-outbreak-news/item/2026-DON615.

14. Publique INdS. Rapport de Situation de la 17eme Epidemie de la Maladie a Virus Ebola/RDC: SitRep No157/MEV B_18/10/2026. Kinshasa: INSP; 2026 18 October 2026.

15. Kelly JD, Hoff NA, Spencer D, Musene K, Bramble MS, McIlwain D, et al. Neurological, Cognitive, and Psychological Findings Among Survivors of Ebola Virus Disease From the 1995 Ebola Outbreak in Kikwit, Democratic Republic of Congo: A Cross-sectional Study. Clin Infect Dis. 2019;68(8):1388–93.

16. Hoff NA, Mukadi P, Doshi RH, Bramble MS, Lu K, Gadoth A, et al. Serologic Markers for Ebolavirus Among Healthcare Workers in the Democratic Republic of the Congo. The Journal of Infectious Diseases. 2019;219(4):517–25.

17. Halbrook M, Merritt S, Hoff NA, Mukadi P, Kompany JP, Musene K, et al. Bundibugyo virus glycoprotein seroreactivity following recombinant vesicular stomatitis virus&#x2013;Zaire Ebola virus glycoprotein vaccination in outbreak-affected populations of the Democratic Republic of the Congo: a longitudinal cohort study. The Lancet. 2026.

18. Merritt S, Mukadi PK, Kompany JP, Halbrook M, Musene K, Martin SA, et al. Detection of Marburg Virus Antibodies 25 Years After Outbreak in Watsa, Democratic Republic of the Congo. The Journal of Infectious Diseases. 2026:jiag155.

19. Myneni RB, Hoffman S, Knyazikhin Y, Privette J, Glassy J, Tian Y, et al. Global products of vegetation leaf area and fraction absorbed PAR from year one of MODIS data. Remote sensing of environment. 2002;83(1-2):214–31.

20. Hansen MC, DeFries RS, Townshend JRG, Sohlberg R, Dimiceli C, Carroll M. Towards an operational MODIS continuous field of percent tree cover algorithm: examples using AVHRR and MODIS data. Remote Sensing of Environment. 2002;83(1):303–19.

21. Long DG, Drinkwater MR, Holt B, Saatchi S, Bertoia C. Global ice and land climate studies using scatterometer image data. Wiley Online Library; 2001.

22. Muyembe J-J, Pan H, Peto R, Diallo A, Touré A, Mbala-Kingebene P, et al. Ebola outbreak response in the DRC with rVSV-ZEBOV-GP ring vaccination. New England Journal of Medicine. 2024;391(24):2327–36.

23. Mulangu S, Alfonso VH, Hoff NA, Doshi RH, Mulembakani P, Kisalu NK, et al. Serologic Evidence of Ebolavirus Infection in a Population With No History of Outbreaks in the Democratic Republic of the Congo. J Infect Dis. 2018;217(4):529–37.

24. Gonzalez JP, Nakoune E, Slenczka W, Vidal P, Morvan JM. Ebola and Marburg virus antibody prevalence in selected populations of the Central African Republic. Microbes and Infection. 2000;2(1):39–44.

25. Hood G, Akoi Bore J, Tipton T, Onivogui Z, Sovogui K, Zoumanigui KZ, et al. Filovirus reactive antibodies in wildlife hunting communities suggest ongoing zoonotic spillover in forested Guinea. Nature Communications. 2026;17(1):9974.

26. Steffen I, Lu K, Yamamoto LK, Hoff NA, Mulembakani P, Wemakoy EO, et al. Serologic Prevalence of Ebola Virus in Equatorial Africa. Emerg Infect Dis. 2019;25(5):911–8.

27. Nyakarahuka L, Schafer IJ, Balinandi S, Mulei S, Tumusiime A, Kyondo J, et al. A retrospective cohort investigation of seroprevalence of Marburg virus and ebolaviruses in two different ecological zones in Uganda. BMC Infect Dis. 2020;20(1):461.

28. Mbala P, Baguelin M, Ngay I, Rosello A, Mulembakani P, Demiris N, et al. Evaluating the frequency of asymptomatic Ebola virus infection. Philos Trans R Soc Lond B Biol Sci. 2017;372(1721).

29. Scotti MT, Kelestemur E, Martin HJ, Novy B, Melo-Filho CC, de Menezes RPB, et al. Conserved Filovirus Proteins as Targets of Broad-Spectrum Antivirals. bioRxiv. 2025.

30. West BR, Moyer CL, King LB, Fusco ML, Milligan JC, Hui S, et al. Structural Basis of Pan-Ebolavirus Neutralization by a Human Antibody against a Conserved, yet Cryptic Epitope. mBio. 2018;9(5).

31. Macneil A, Reed Z, Rollin PE. Serologic cross-reactivity of human IgM and IgG antibodies to five species of Ebola virus. PLoS Negl Trop Dis. 2011;5(6):e1175.

32. Flyak AI, Shen X, Murin CD, Turner HL, David JA, Fusco ML, et al. Cross-Reactive and Potent Neutralizing Antibody Responses in Human Survivors of Natural Ebolavirus Infection. Cell. 2016;164(3):392–405.

33. Negredo A, Palacios G, Vázquez-Morón S, González F, Dopazo H, Molero F, et al. Discovery of an ebolavirus-like filovirus in europe. PLoS Pathog. 2011;7(10):e1002304.

34. Yang XL, Tan CW, Anderson DE, Jiang RD, Li B, Zhang W, et al. Characterization of a filovirus (Měnglà virus) from Rousettus bats in China. Nat Microbiol. 2019;4(3):390–5.

35. Wight J, Schulz H, Banadyga L. Antibodies cross-reactive with Bundibugyo virus in ferrets vaccinated with Ebola virus vaccine. Emerging Infectious Diseases. 2026;32(8):1360.

36. Lhomme E, Wiedemann A, Ayouba A, Ben-Farhat S, Thaurignac G, Roy C, et al. Cross-Reactive Bundibugyo Antibody Responses after Receipt of Licensed Ebola Vaccines. New England Journal of Medicine. 2026.

37. Wight J, Liu G, Chan M, Medina SJ, Lu D, Cao W, et al. rVSV-EBOV vaccination protects ferrets from lethal Bundibugyo virus disease. bioRxiv. 2026:2026.08.24.746878.

38. R&D Blueprint (RDB) TAGoCVPT-C. Third meeting of the WHO Technical Advisory Group on candidate vaccine prioritization (TAG-CVP) for Bundibugyo virus disease outbreak response: meeting report, 31 July 2026. 2026. Report No.: WHO Reference Number: B09865.

39. Lu LL, Suscovich TJ, Fortune SM, Alter G. Beyond binding: antibody effector functions in infectious diseases. Nat Rev Immunol. 2018;18(1):46–61.

40. Ghosh K, Javeri KN, Mohanty D, Parmar BD, Surati RR, Joshi SH. False-positive serological tests in acute malaria. Br J Biomed Sci. 2001;58(1):20–3.

41. Shaffer KCL, Hui S, Bratcher A, King LB, Mutombe R, Kavira N, et al. Pan-ebolavirus serology study of healthcare workers in the Mbandaka Health Region, Democratic Republic of the Congo. PLOS Neglected Tropical Diseases. 2022;16(3):e0010167.

42. Bangelesa F, Akilimali P, Hosner R, Buckee C, Shoemaker TR, Kaba D. Population mobility and Ebola case containment in Ituri, DR Congo. The Lancet. 2026.

